# GraftIQ: A Hybrid Multi-Class Neural Network Integrating Clinical Insight for Multi-Outcome Prediction in Liver Transplant Recipients

**DOI:** 10.1101/2024.10.28.24316280

**Authors:** Divya Sharma, Neta Gotlieb, Daljeet Chahal, Sara Naimimohasses, Ankit Ray, Yoojin Han, Sara Gehlaut, Maryam Shojaee, Surabie Sivanendran, Maryam Naghibzadeh, Amirhossein Azhie, Sareh Keshavarzi, Kai Duan, Leslie Lilly, Nazia Selzner, Cynthia Tsien, Elmar Jaeckel, Wei Xu, Mamatha Bhat

## Abstract

**Background and Aims:** Liver transplant recipients (LTRs) are at risk of developing graft injury, leading to cirrhosis and reduced survival. Liver biopsy remains the gold standard method for the diagnosis of graft pathology but is invasive and risky. Our study aimed to develop a novel hybrid multi-class neural network (NN) model ‘GraftIQ’ integrating clinician expertise for non-invasive diagnosis of graft pathology.

**Methods:** Graft injury diagnosis was based on liver biopsies from LTRs (1992-2020). Demographic, clinical, and laboratory data from the 30 days before biopsy were used to train a multi-class NN model to classify biopsies into six categories. The dataset was split into 70% training and 30% test sets, with external validation on additional biopsies from 2020-2024. To enhance predictive capabilities, clinician expertise was integrated with neural network predictions using Bayesian fusion to combine clinician-provided probabilities with data-driven outcomes.

**Results:** Our dataset comprises 5,217 biopsies categorized into six graft etiology groups. In response to findings from expert versus machine implementation analysis, Bayesian fusion of clinical expertise and NN predictions enhanced predictive performance. GraftIQ (MulticlassNN + clinical insight) achieved an overall AUC of 0.902 (95% CI: 0.884, 0.919), improving from an AUC of 0.885 using the NN alone. Robustness validated through 10-fold internal cross-validation and external validation, showed AUC improvements of 10-16% compared to conventional machine learning approaches.

**Conclusion:** Our multi-class neural network model demonstrates high accuracy in predicting common causes of graft pathology. Through the integration of clinician expertise, we observed an improvement in its performance, affirming the effectiveness of GraftIQ as a valuable clinical decision support tool.

**Availability of code:** https://github.com/divya031090/multiclassNN

## INTRODUCTION

Liver transplantation (LT) is a life-saving measure for selected patients with end stage liver disease ^1^. Despite tremendous improvement in LT outcomes over recent decades, liver transplant recipients (LTRs) remain at risk of developing graft injury of various etiologies. Graft injury can result in fibrosis and cirrhosis over time, potentially resulting in graft loss in 25% of LTRs ^2^. Graft viability after LT is dependent on the prompt recognition of post-transplant pathologies. Promptly starting treatments like high-dose steroids for rejection or antiviral therapy relies on identifying the cause of graft injury to prevent long-term dysfunction.

Graft injury is often suspected via elevated liver enzymes during routine bloodwork. However, biochemical tests are non-specific and it is difficult to establish a cause of injury based on these alone ^3^. Liver biopsy has therefore remained the gold standard for the diagnosis of graft pathology^4^. In fact, with improvements in post-transplant survival over the last 2 decades, repeat evaluations of graft function via biopsy have become more frequent^5^. Liver biopsy is subject to sampling error as well as complications such as bleeding, infection^6^ and is often unavailable in a timely manner. Time constraints mean that hepatologists often have to make empiric decisions before liver biopsy is carried out. Such decisions are based on clinical data (age, indication for transplant, time after transplant, diabetes, obesity, immunosuppression regimen) and liver biochemical patterns. Thus, there is a clinical need to develop reliable, non-invasive methodologies that can establish or rule out specific etiologies of graft injury prior to biopsy, allowing rational therapeutic decisions to be made as quickly as possible.

Machine learning (ML), specifically Neural Networks (NN) are efficient in analyzing large, complex, and heterogeneous datasets, generating reproducible predictions and classifications on previously unseen data^7^. Previous studies have demonstrated the feasibility of convolutional NNs to generate accurate prognostic predictions and fibrosis detection in various chronic liver diseases by leveraging blood test patterns and interrelationships between variables^8,9^. When applied to liver transplantation, ML has emerged as a promising methodology to stratify patient risk and predict post-transplant outcomes^10-12,30^. Convolutional NNs have been applied to help predict waitlist mortality^13^, donor-recipient matching^14,15^ and HCC recurrence^16^. While ML models have the potential to process and analyze vast amounts of data quickly and efficiently, it is important to recognize that ML models may lack the nuanced understanding, and contextual knowledge that experienced clinicians bring to patient care.

In this study, we hypothesized that combining the extensive, prior knowledge of causal and correlational associations that human experts possess with a machine-learned model would increase model generalizability. To address the same, we have developed a novel hybrid multiclass neural network model ‘GraftIQ’, that aims to predict the etiology of a graft injury using richly annotated clinical, demographic and laboratory data from LT recipients. As opposed to neural networks being commonly used for binary classification, the novelty of our approach lies in its multi-class classification of 6 diagnostic categories. This enables our approach to be a one-step methodology for multicategory classification through a single comprehensive model. Beyond this, another novelty of our methodology is the Bayesian fusion-based integration of clinician feedback to refine predictions of our ML model for being applicable in the clinical setting. Our hybrid ML tool ‘GraftIQ’ could potentially reduce dependence on longitudinal liver biopsies and lead to earlier therapeutic interventions, improving graft viability and patient survival over time.

## MATERIALS and METHODS

### 1. Data collection and setting

Demographic, clinical and laboratory data of all adult LTRs having undergone liver biopsies between January 17^th^, 1992 and June 16^th^, 2020 at the Ajmera transplant center, UHN, Toronto, Canada form the main dataset of our study. This study was approved by the Research Ethics Board at UHN (REB study # 21-6170). Since data was retrieved from medical records, exemption from informed consent was granted by the REB committee.

## 2. Study Design

### 2.1 Definition and diagnosis of post-transplant complications

The first part of the study was to establish the most common etiologies for graft injury in LTRs from available biopsies. Biopsies were reviewed by two separate reviewers and labeled according to the appropriate diagnosis from the pathologist’s biopsy report. Biopsies were labeled as normal, acute cellular rejection (ACR), antibody-mediated rejection, biliary obstruction (BO), congestion, alloimmune hepatitis (AIH), viral (Hepatitis C, Hepatitis B, Cytomegalovirus (CMV), Epstein Barr virus (EBV)), metabolic-associated steatohepatitis (MASH) and toxic/drug-induced graft injury. Dual diagnoses were excluded from the analysis. The categories that were considered for statistical and ML analysis included ACR, BO, AIH, Hepatitis C infection (HCV), congestion, and MASH. The biopsies that read as normal as well as all other remaining diagnoses were grouped together as ‘Others’ due to the small number of samples as shown in Figure 1(b).

**Figure 1:**
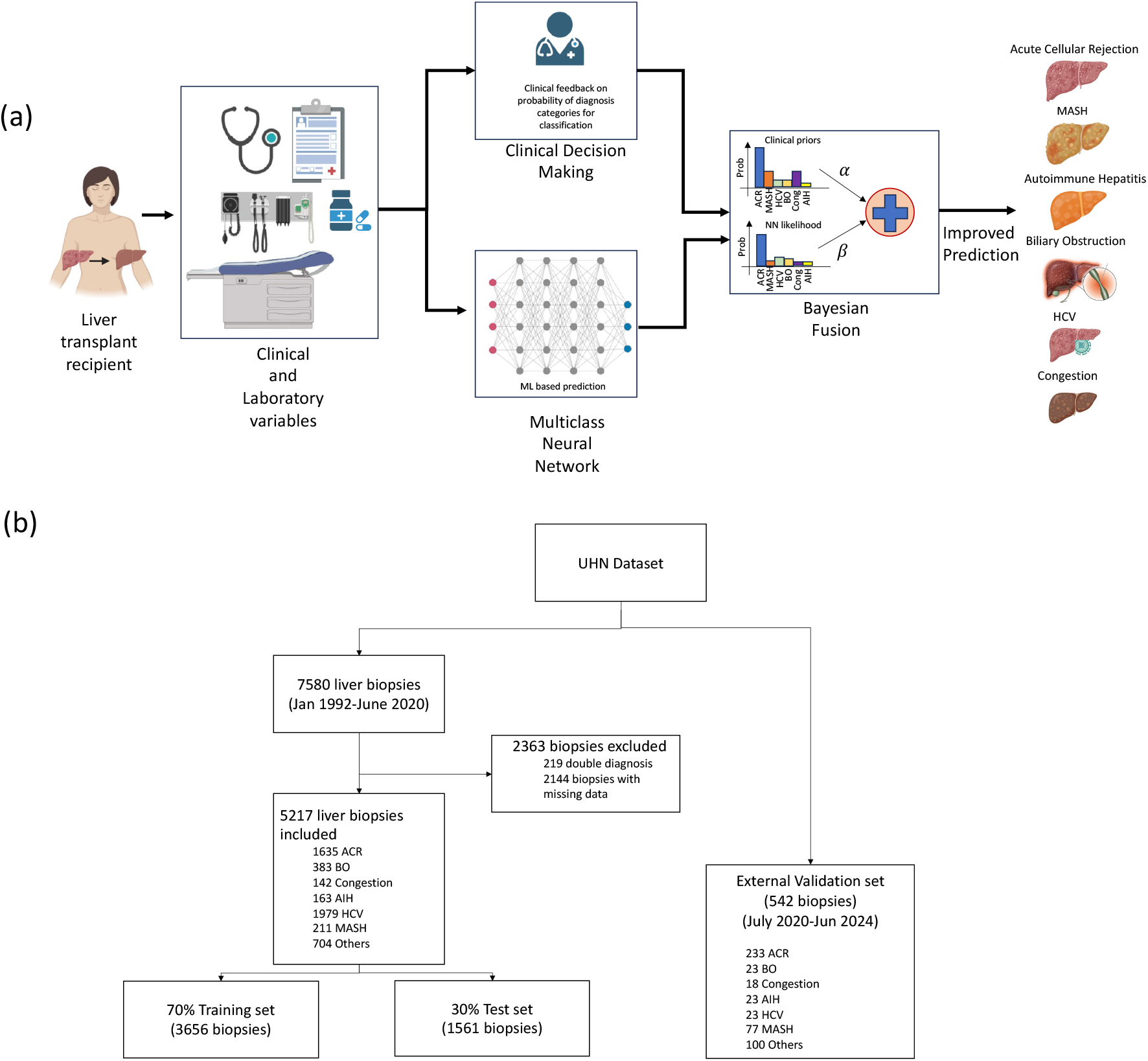
Overall Framework and Study Design. (a) Schematic representation of our hybrid multiclass neural network, ‘Graft IQ,’ which combines clinician expertise with multiclass neural network capabilities to predict the cause of graft injury. (b) Flowchart detailing study design and data distribution

### 2.2 Demographic and clinical data for each diagnosis

The next step was to allocate demographic and clinical variables measured closest to the biopsy date, up to 30 days before each biopsy. The data included Aspartate aminotransferase (AST), Alanine aminotransferase (ALT), Alkaline phosphatase (ALP), bilirubin, international normalized ratio (INR), white blood cells, hemoglobin, platelets, tacrolimus, and cyclosporine levels. Each biopsy was considered a ‘subject’, and the data was considered for each biopsy separately with an aim to assess which clinical variables triggered the biopsy. Demographic and pre-transplant variables (transplant indication, Model for End-Stage Liver Disease [MELD], donor type) for each biopsy were included in the analyses, as well as clinical data prior to the biopsy date (cholangitis, body mass index [BMI], diabetes, hypertension, and dyslipidemia). Missing data if any was imputed using mean imputation method in MICE library in R (Missing data details in Supplementary Table 6).

### 2.3 Implementation Analysis

In this implementation analysis, 12 hepatologists with diverse expertise were chosen to compare their predictive abilities with GraftIQ’s algorithm. Using a dataset of 30 cases covering six pathology categories, diagnostic accuracy was evaluated by comparing the ML model’s predictions to independent diagnoses by hepatologists. Overall accuracy was assessed to compare the ML model’s predictions with the independent diagnoses made by the hepatologists. This analysis helped us obtain insights into how the hepatologists made the prediction and narrow down on 6 simple clinical rules that hepatologists use to distinguish between etiologies namely: for BO, ALP and bilirubin should be high; for ACR, age of graft should be low given the increased risk in the early post-transplant phase; ALT>ALP and immune-mediated liver disease as an indication for transplant help to distinguish alloimmune hepatitis. For MASH, the age of graft should be higher given its progressive nature, in addition to metabolic risk factors, including an elevated BMI in conjunction with more modest elevations of ALT. For HCV, ALT>AST with HCV as an indication for transplant and the presence of positive HCV serology are considered. Finally for congestion, ALP and INR should be high without significant elevations of bilirubin in addition to the age of graft being low. These rules were additionally used in our clinical integration step.

### 2.4 Machine learning analysis

#### 2.4.1 Multiclass Neural Network model

We propose a Neural Network model with multiple classes to carry out the classification task (Figure 1(a)). Usually, ML classification algorithms restrict the possible outcomes to one of two values (a binary, or two-class model), however, given that our outcome included multiple primary diagnosis categories, we modified the learning function in the neural network to predict multi-class output. In our neural network methodology, we adopt the softmax approach, a multinomial logistic regression extension that directly supports multi-class classification. In our case, with six different diagnosis categories, the output layer consists of six nodes, each representing one of the classes. The softmax activation function is employed for each node, producing a probability distribution across all classes. The model is trained using the categorical cross-entropy loss function, which is well-suited for multi-class scenarios. This methodology fosters an ensemble-like behavior within the neural network, allowing it to collectively predict all classes while maintaining interpretability and computational efficiency.

We divided our dataset into 70% training and 30% test set for model evaluation. Internal 10 times 10-fold cross-validation was performed to tune hyperparameters and compare our multiclass NN approach with other conventional ML approaches namely, random forest, support vector machines, logistic, lasso and ridge regression (hyperparameter optimization details provided in Supplementary Tables 4-5). For external validation of the model, we collected an additional 542 liver biopsies and clinical data from the UHN database between July 2020 until June 2024. Similar to the main dataset, two separate reviewers labeled the biopsies according to the appropriate diagnosis from the pathologist’s biopsy report and divided them into the same seven categories (ACR, AIH, congestion, BO, HCV, MASH and others). To mitigate the potential impact of the relatively small sample size in some of the categories, we employed a repeated bootstrapping approach to assess the robustness and generalizability of the model. Specifically, we generated 1,000 bootstrap samples from the external validation set, each consisting of a random sample with replacement. The model’s performance was evaluated using mean AUC and 95% confidence intervals to estimate the model’s stability and ensure it was not overfitting.

#### 2.4.2 Expert-Enhanced Adaptive Integration

To enhance the predictive capabilities of the neural network, we introduced a novel approach for integrating clinician expertise into the posterior probability calculation. Clinician input was obtained in the form of probability assessments for each graft injury category based on the 6 clinical rules mentioned in Section 2.3. In this approach, if a clinical rule is satisfied for a specific diagnostic category, the probability for that category is set to 1. If multiple clinical rules are satisfied, the probabilities for each corresponding diagnostic category are distributed equally, ensuring that their sum equals 1. These clinician-provided probabilities of diagnosis categories were encoded as prior knowledge, reflecting expert assessments of diagnosis likelihoods based on patient data and to guide the inference process. Concurrently, a neural network architecture as illustrated in section 2.4.1, was employed to compute the likelihood of each diagnosis category from observed data. Bayesian inference principles were then applied to fuse the prior knowledge provided by clinicians with the likelihood computed by the neural network. This Bayesian fusion process yielded a posterior probability distribution over diagnosis categories, capturing the integration of clinical expertise and data-driven predictions. Subsequently, value-based probabilistic inference techniques were employed to make decisions based on the posterior probability distribution in the last layer of the neural network. The integration was achieved through a weighted combination of the probabilities generated by the machine learning model and the clinician. The posterior probability P_integrated_(C_i_) for each diagnosis category C_i_ was calculated using the following

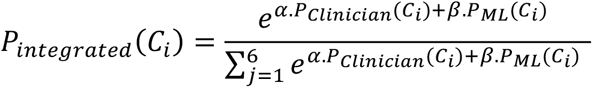

Where, *P*_*ML*_(*C*_*i*_) represents the probability assigned by the multi class NN ML model, *P*_cl*i*n*i*c*i*an_(*C*_*i*_) is the probability provided by the clinician for category C_i_ and *α* and *β* are the weight parameters to assign confidence clinician prediction and the ML prediction respectively.

This iterative feedback loop not only provides valuable insights into clinicians’ domain expertise but also empowers the ML model to continuously learn from real-world scenarios, potentially resulting in more precise predictions.

#### 2.4.3 Extract Important Features through Neural Networks

To identify the important variables in our predictive modeling, we used the Integrated Gradient (IG) methodology which is an interpretability technique for deep neural networks^17^. We calculated gradients to measure the relationship between changes to a variable and corresponding changes in the model’s predictions. The gradient informs which variable has the strongest effect on the model’s predicted class probabilities where the higher the gradient, the more important the feature is considered to the classification task.

## RESULTS

### 1. Patient population

A total of 1791 patients were identified for analysis. Mean recipient age was 52.4 ± 11.0 years, and mean donor age was 43.8 ± 16.5 years. A total of 601 patients (34%) were female, and 1190 (66%) male. Mean recipient weight was 79.2 ± 17.9 kilograms, and 388 patients (27%) had BMI over 30. Comorbidities included diabetes in 282 patients (16%), hypertension in 237 patients (14%) and dyslipidemia in 59 patients (3%). Mean MELD at time of transplantation was 18.3 ± 9.3. A total of 448 patients (25%) received living donor liver transplant, and 1343 (75%) received deceased donor liver transplant. 90 patients (5%) developed recurrent hepatocellular carcinoma (HCC) and 138 patients (8%) developed cholangitis. The indications for transplant are described in Supplementary Table 1. The most common indication was Hepatitis C with 711 biopsies (40%), followed by immune-mediated liver diseases (AIH, Primary biliary cholangitis, Primary sclerosing cholangitis) at 289 (17%), alcohol-related liver disease at 211 (12%) and MASH at 121 (7%).

### 2. Disease cohort characteristics

7580 liver biopsies were available from our post-transplant database. After careful review and exclusion of biopsies with missing data and double diagnoses, a total of 5217 biopsies remained and were included in the analysis. This total is higher than the total number of patients, as many patients had more than 1 biopsy over their post-transplant course. From this total, we identified the diagnostic categories of ACR, AIH, BO, congestion, HCV, MASH, and others but focused on the first six for our analysis. A total of 1979 biopsies were consistent with HCV, 1635 with ACR, 383 with BO, 211 with MASH, 163 with AIH, 142 with hepatic congestion, and 704 considered as others. We documented mean values of laboratory variables at time of biopsy and up to 30 days prior for each category. Details about these features for each disease cohort provided in Table 1.

**Table 1:** Clinical and demographic features for study groups. Presented as mean (standard deviation) unless otherwise specified.

|  | ACR | AIH | BO | Congestion | HCV | MASH |
| --- | --- | --- | --- | --- | --- | --- |
| Biopsies (N) | 1635 | 163 | 383 | 142 | 1979 | 211 |
| Recipient age (years) | 50.5 (11.4) | 49.8 (12.2) | 51.2 (11.8) | 47.2 (12.3) | 54.2 (8.3) | 56.2 (8.9) |
| Donor age (years) | 43.1 (16.9) | 43.2 (17.7) | 43.6 (14.0) | 42.8 (21.8) | 44.1 (17.8) | 43.5 (19.5) |
| Weight (kg) | 76.8 (16.4) | 77.0 (17.1) | 80.4 (17.5) | 74.1 (13.3) | 80.4 (17.0) | 89.1 (20.0) |
| BMI (kg/m<sup>2</sup>) | 26.8 (4.8) | 27.2 (6.0) | 27.7 (5.6) | 26.0 (2.7) | 27.7 (4.8) | 29.9 (6.0) |
| ALT (IU/L) | 218.52<br>(232.72) | 154.64<br>(160.04) | 135.73<br>(179.93) | 100.39<br>(104.79) | 115.01<br>(128.51) | 116.21<br>(110.61) |
| AST (IU/L) | 138.14<br>(185.04) | 133.71<br>(145.71) | 123.44<br>(487.64) | 91.53<br>(162.33) | 98.56<br>(107.16) | 110.84<br>(106.34) |
| ALP (IU/L) | 376.42<br>(333.82) | 234.21<br>(201.01) | 397.69<br>(315.69) | 342.22<br>(322.42) | 170.63<br>(140.53) | 201.82<br>(210.52) |
| WBC (10<sup>9</sup>/L) | 8.30 (5.10) | 6.19 (3.49) | 8.42 (8.22) | 7.47 (4.87) | 5.90 (2.70) | 6.63 (3.33) |
| Bilirubin (μmol/L) | 67.56 (93.06) | 36.19<br>(72.49) | 106.29<br>(119.79) | 59.25<br>(106.75) | 23.18<br>(40.18) | 34.36 (61.96) |
| Creatinine (μmol/L) | 104.37<br>(54.97) | 116.27<br>(93.77) | 118.48<br>(82.38) | 117.30<br>(56.90) | 117.75<br>(60.75) | 111.64<br>(70.94) |
| HGB (g/L) | 110.27<br>(20.57) | 129.10<br>(20.60) | 106.83<br>(21.73) | 111.26<br>(23.16) | 129.47<br>(19.57) | 133.19<br>(22.59) |
| MELD at Time of OLT | 13 (6) | 11 (5) | 16 (8) | 13 (7) | 11 (4) | 11 (5) |
| Sodium (mmol/L) | 138.12 (4.22) | 139.45<br>(4.15) | 137.65<br>(4.75) | 139.06<br>(5.06) | 139.71<br>(3.31) | 139.26 (4.26) |
| Platelets (10<sup>9</sup>/L) | 185.57<br>(108.37) | 162.85<br>(80.85) | 178.81<br>(102.01) | 175.63<br>(111.93) | 170.11<br>(79.81) | 178.66<br>(75.66) |
| Albumin (μmol/L) | 33.60 (7.60) | 37.12 (6.32) | 32.42 (7.32) | 33.86 (8.16) | 38.37 (5.77) | 39.15 (7.35) |
| INR | 1.73 (0.83) | 1.33 (0.43) | 1.41 (0.61) | 1.52 (0.72) | 1.19 (0.29) | 1.84 (1.04) |
| Tacrolimus (ng/mL) | 10.06 (5.66) | NA | 9.11 (0.61) | NA | 9.28 (5.08) | 9.36 (5.06) |
| Cyclosporine (ng/mL) | 356.12<br>(374.62) | 741.89<br>(155.29) | 322.70<br>(266.90) | 290.66<br>(255.86) | 395.74<br>(294.84) | 364.64<br>(262.44) |

### 3. Results of implementation analysis

As shown in Figure 2, for the 30 cases chosen for the implementation analysis, the ML model exhibited higher predictive accuracies, surpassing hepatologists in every evaluated category. Notably, the ML tool achieved a perfect 100% accuracy for Alloimmune hepatitis, BO, HCV, MASH, showcasing its robust diagnostic capabilities. In contrast, hepatologists demonstrated comparatively lower accuracy rates, particularly in predicting ACR, BO, congestion, and MASH. The instances where our ML model misclassified ACR (67%) and congestion (80%) categories shed light on the importance of integrating clinical expertise into our predictive framework. In the case of ACR, cases were misclassified as MASH, despite elevated liver enzymes and blood tests, as the ML model failed to consider the patient’s low age of graft, a key indicator against MASH. Similarly, in congestion misclassified as HCV, the model overlooked the absence of HCV as an indication for transplant. These discrepancies underscore the necessity of incorporating clinical insights to align more closely with the complexities of real-world clinical scenarios.

**Figure 2:**
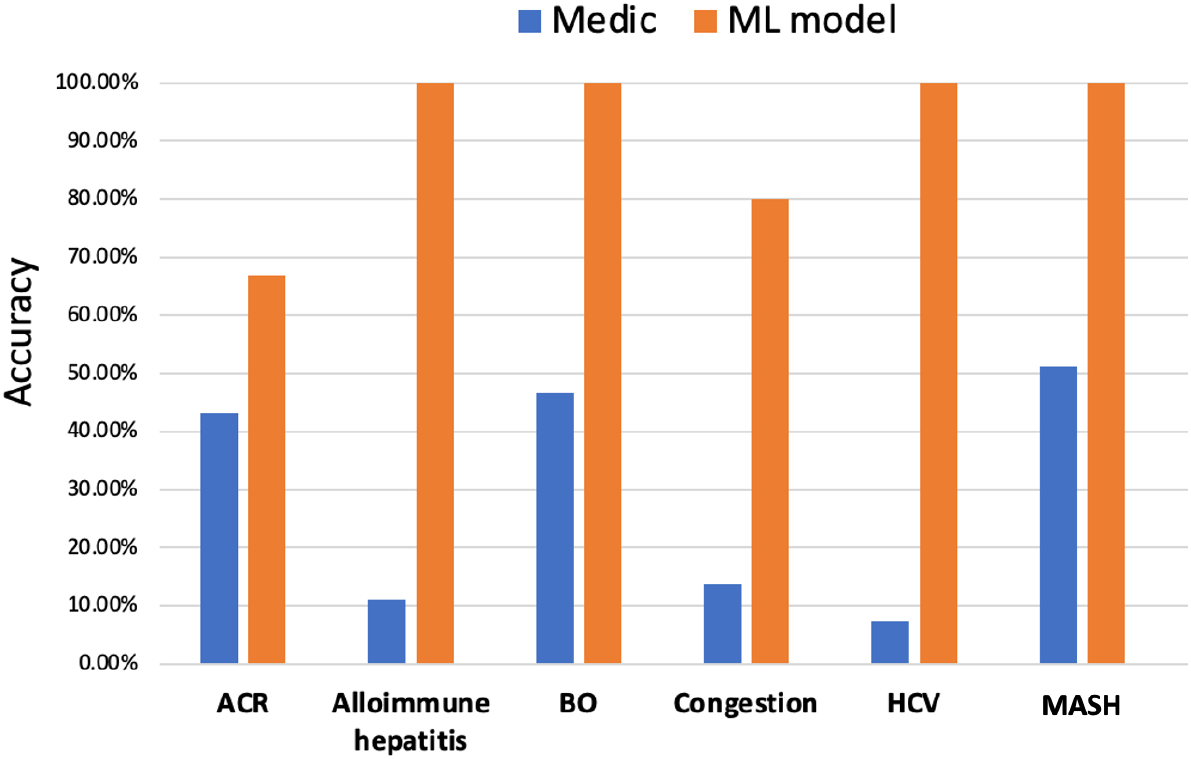
Expert vs Machine Implementation analysis. Comparison of multiclass NN based ML model’s prediction using clinical and lab data vs averaged prediction accuracy provided by 12 hepatologists (medics)

### 4. Predictive Performance evaluation

#### 4.1 Results on the test set

##### 4.1.1 Utilizing multiclass Neural Network model standalone

Firstly, we evaluated the performance of our multiclass NN based ML model independently, without integrating any clinical expertise for predicting each diagnosis category as shown in Table 2. The best performance was obtained for MASH post-transplant complications with Area Under the Curve (AUC) of 0.929 calculated using the Receiver Operating Characteristic (ROC) curve with a sensitivity, and specificity of 0.89 and 0.92 respectively, followed by AIH and congestion, with AUC of 0.924 and 0.922 respectively. The overall AUC was obtained by averaging the AUC obtained for each individual category. In our case, the overall AUC for our neural network methodology on the test set was obtained to be 0.885 [95% Confidence Interval (C.I.): 0.864, 0.901].

**Table 2:**
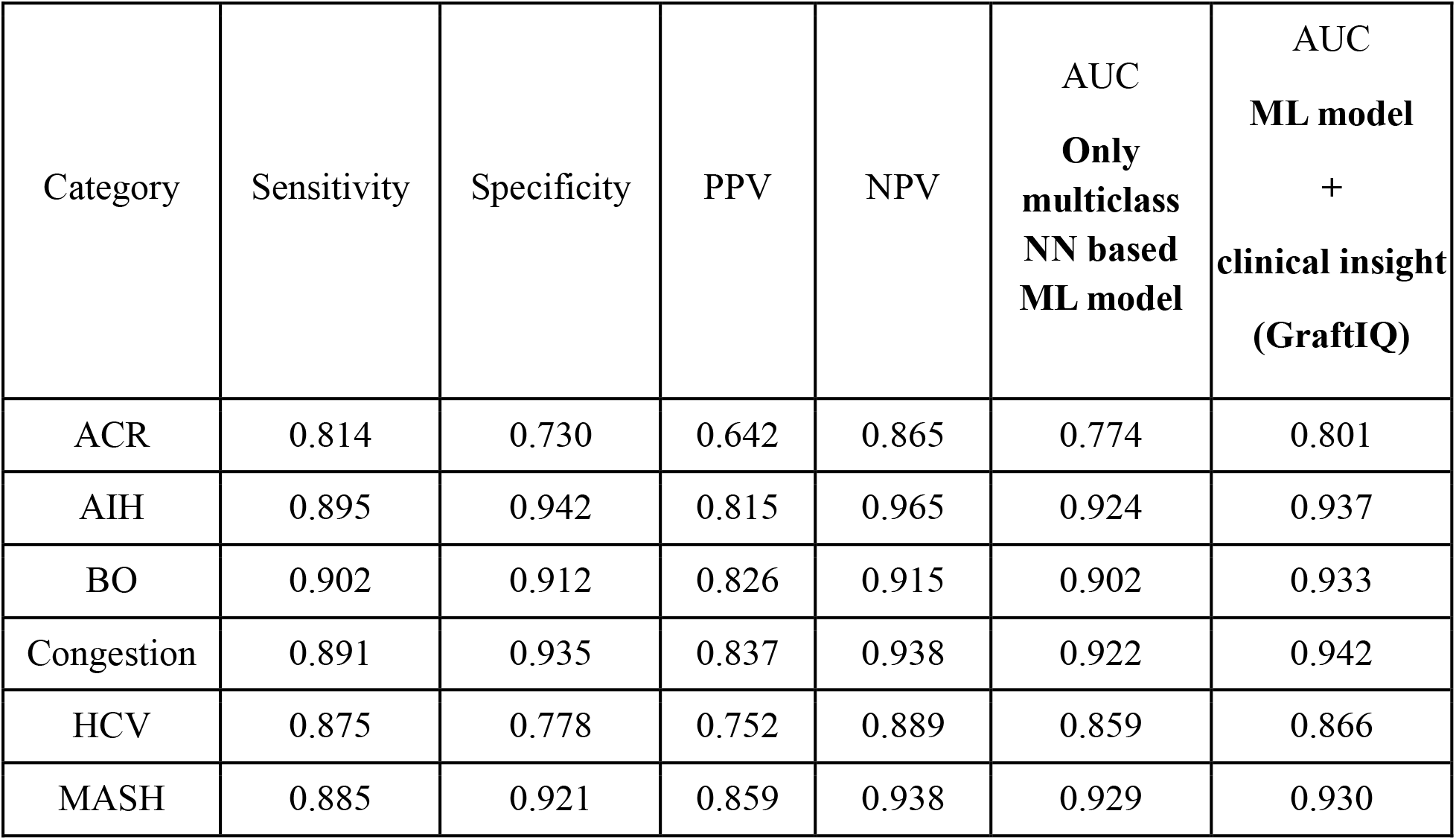
Performance metrics obtained through evaluation of the hybrid neural network algorithm ‘GraftIQ’ in predicting diagnosis categories. Performance metrics obtained using only the multiclass NN as well as the hybrid neural network algorithm (multiclass NN + clinical insight) for each complication group. Comprehensive analysis in terms of Sensitivity, Specificity, Positive Predictive value (PPV), Negative Predictive Value (NPV) and Area Under the curve (AUC) is represented for each category. The higher each metric, the better the classification is. Note the improvement in AUC values as observed upon integrating clinician expertise into the neural network model for each diagnosis category.

##### 4.1.2 Utilizing “GraftIQ”, hybrid model integrating NN prediction and clinical insight

As shown in Table 2, column 7, the incorporation of clinician-based probabilities (with *α* = 0.2 and *β* =0.8 for fusion after tuning as shown in Supplementary Table 2) into the final layer of our neural network model resulted in improvement in predictive performance for each diagnosis category. Specifically, the AUC values surpassed 0.8 for every category (notably high improvement for ACR prediction), representing a significant improvement compared to predictions made using the ML model alone. The overall AUC based on integrating clinical expertise improved from 0.885 to 0.902.

We then compared our neural network model to other conventional machine learning models (refer Table 3) and found that the neural network performs better in terms of overall AUC for classification with AUC of 0.902, 95% C.I.: [0.884, 0.919] as compared to the second-best approach Random Forest with an AUC of 0.823 [95% C.I.: [0.812, 0.839]. Regression approaches performed relatively less accurately in terms of multi-class classification with Logistic Regression with an AUC of 0.767 [95% C.I.: 0.626, 0.796], Lasso with an AUC of 0.783 [95% C.I.: 0.769, 0.802] and Ridge regression with an AUC of 0.781 [95% C.I.: 0.771, 0.811] justifying the prominence of Neural Networks in understanding the non-linear relationships in the data as well as in assigning subjects accurately to one of the multiple categories of diagnosis.

**Table 3:**
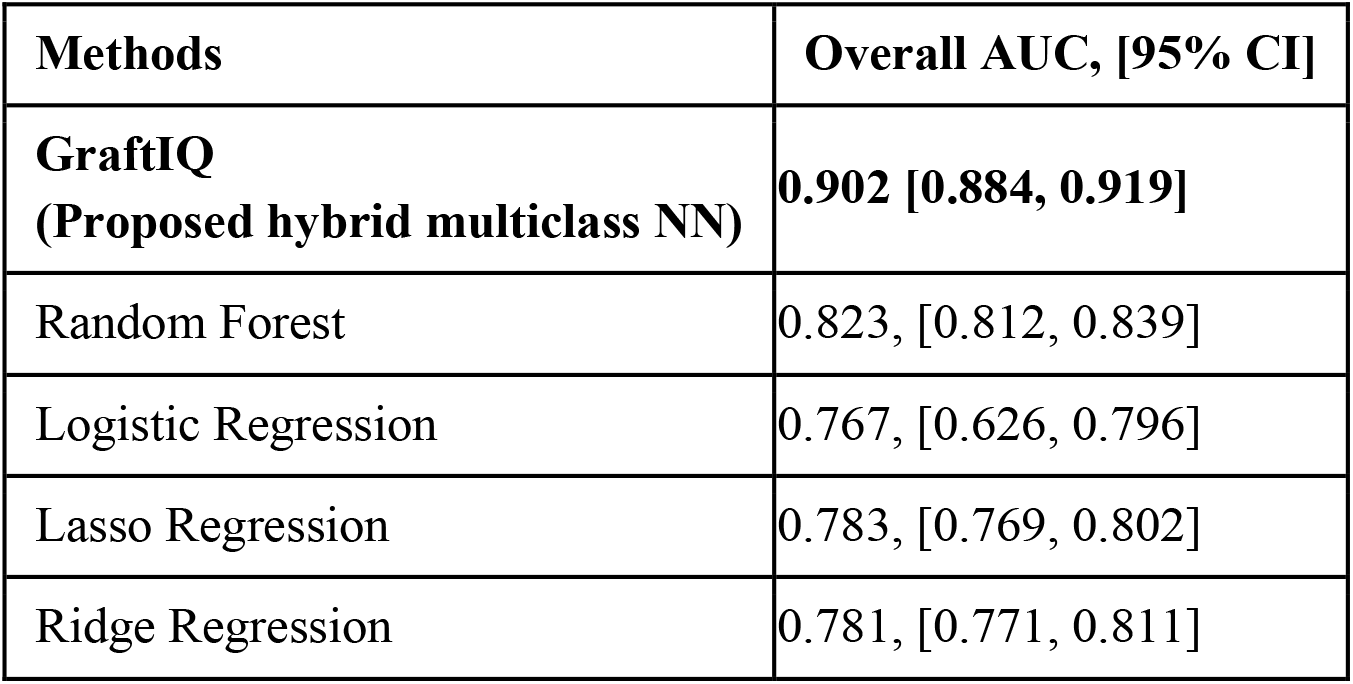
Comparative analysis of proposed hybrid GraftIQ model vs. conventional Machine Learning algorithms for comparing overall AUC averaged across six diagnosis categories.

| Methods | Overall AUC, [95% CI] |
| --- | --- |
| **GraftIQ**<br>**(Proposed hybrid multiclass NN)** | **0.902 [0.884, 0.919]** |
| Random Forest | 0.823, [0.812, 0.839] |
| Logistic Regression | 0.767, [0.626, 0.796] |
| Lasso Regression | 0.783, [0.769, 0.802] |
| Ridge Regression | 0.781, [0.771, 0.811] |

To make our neural network methodology more explainable and clinically relevant, we also computed the variable importance of each clinical feature in the classification task for individual categories. The higher the gradient obtained through the Integrated Gradient methodology detailed in subsection 2.4.3, the more important the feature is towards the classification task. As shown in Figure 3, ALT, ALP and hemoglobin were the top 3 features important to the classification of subjects in the ACR category. Similar plots for the rest of the five diagnosis categories are provided in the Supplementary document (Supplementary Figures 1-5).

**Figure 3:**
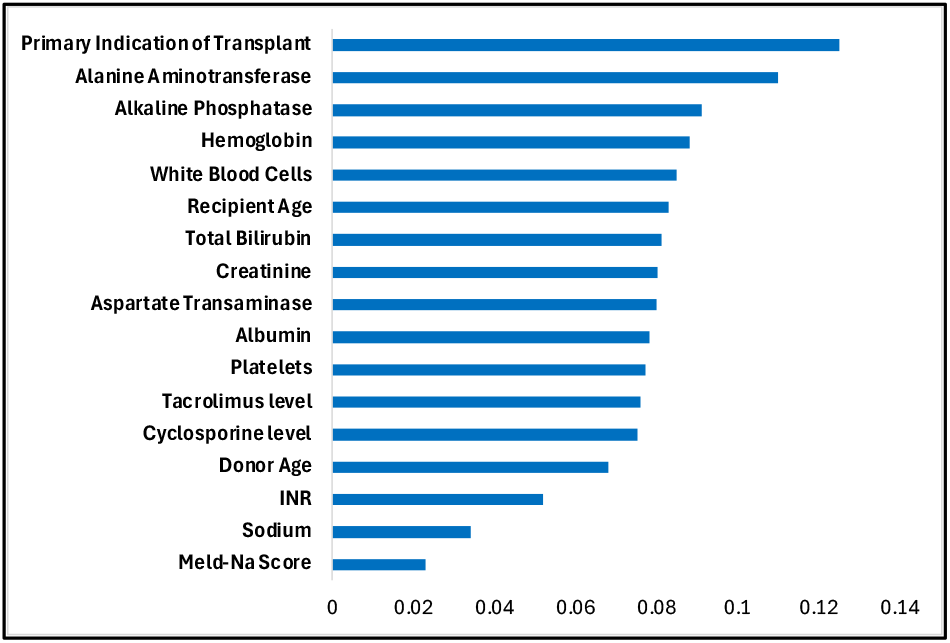
Importance plot ranking features relevant to classification of subjects into the Acute Cellular Rejection (ACR) category. The x-axis represents the gradient in neural network learning. The higher the gradient, the more important the feature is.

#### 4.2 Results on external validation set

542 biopsies were reviewed and divided according to the 6 relevant categories along with other biopsies. 233 biopsies were consistent with ACR, 68 with biliary obstruction, 77 with MASH, 18 with congestion, 23 with HCV, 23 with AIH and 100 considered as others. We focused on the first six categories for clinical significance. The model performance in the external test set was in line with our main results with the best performance obtained for AIH with mean AUC of 0.962 followed by MASH and BO (Table 4). The overall AUC by averaging the AUCs obtained for each individual category was 0.934 [95% Confidence Interval (C.I.): 0.909, 0.959] showing the robustness of our methodology on a completely unseen external validation set.

**Table 4:**
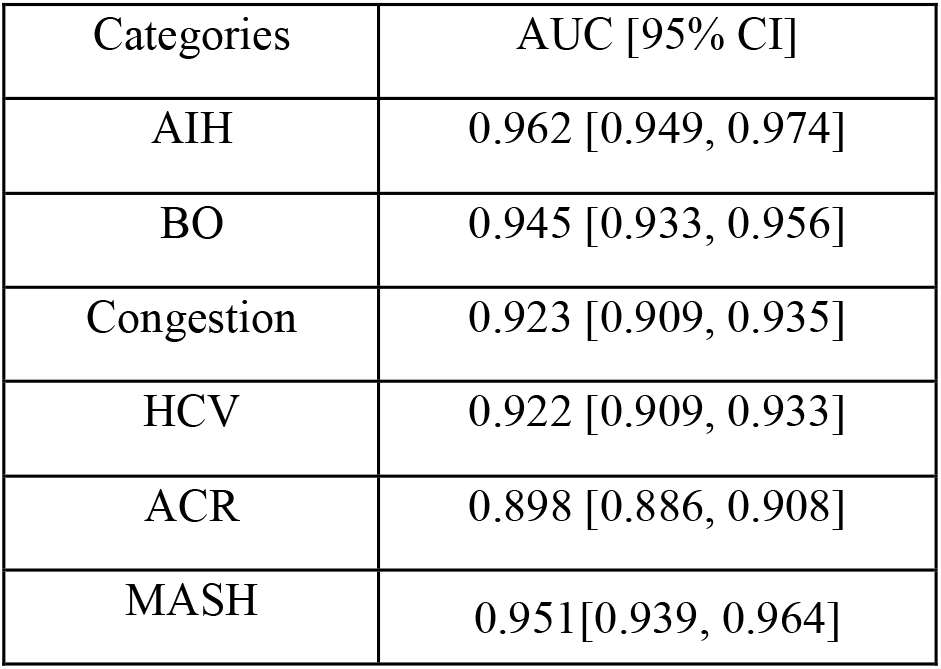
Evaluation results using mean Area Under the Curve (AUC) metric and 95% Confidence Intervals (CI) on each diagnosis category in the external validation dataset using the proposed GraftIQ model.

### 5. Demonstration of Clinical Relevance

To demonstrate further clinical relevance of GraftIQ, we randomly chose one patient from each diagnostic category and applied the algorithm to obtain the probability of each possible diagnosis. We then manually reviewed the raw lab values for each patient to determine if the ML output made clinical sense or provided an expedited path to diagnosis that would otherwise have required further investigation.

Graphs demonstrating the probability of each diagnosis for each selected patient are displayed in Figure 4(a). For example, in patient 1147 with a liver biopsy demonstrating ACR, our algorithm determined an 81% probability of a diagnosis of ACR, followed by a 6% probability of MASH, 5% probability of HCV, 4% probability of BO, 3% probability of AIH and 1% probability of congestion. Subsequent review of the lab parameters for this patient that the algorithm used as part of its analysis demonstrated an ALP of 803, total bilirubin of 147, ALT 227, AST of 141.

**Figure 4:**
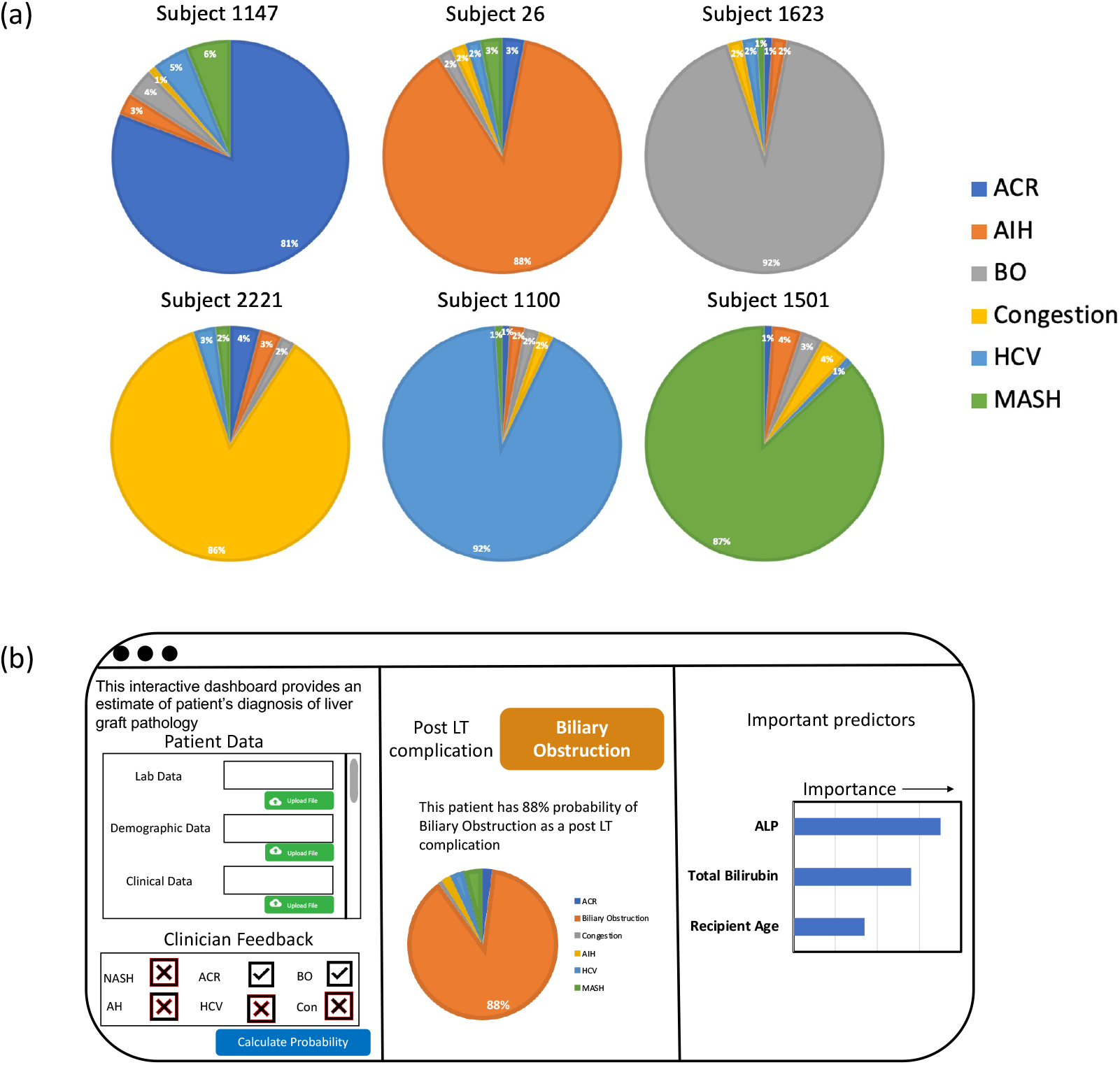
Clinical relevance and integration of GraftIQ. (a) Pie charts illustrating probability of a subject being classified into one of the six diagnosis categories using hybrid GraftIQ model. Example subjects selected based on their true diagnosis categories in the dataset, and higher probability in the pie chart represents their predicted class, e.g., for Subject 1501, the true diagnostic category in the test set was MASH and our hybrid model predicted subject being classified as MASH with a probability of 87%. (b) Illustration of a proposed interactive clinician-facing dashboard for clinical integration of our GraftIQ model.

For recurrent AIH, our hybrid model demonstrated an 88% probability of AIH. Review of the labs demonstrated ALP of 617, ALT of 377, AST of 377 and total bilirubin of 82. For post-transplant congestion, ALP was 331, ALT 66, AST 46, and bilirubin 25, whereas for HCV, ALP was 328, ALT 122, AST 66 and bilirubin was 15. Again, GraftIQ was able to identify these diagnoses with probabilities of 86% and 92%, respectively. Our patient with recurrent MASH had an ALP of 86, ALT 109, AST 35 and our algorithm identified this diagnosis with a probability of 87%.

As can be seen from these results, the pattern of liver tests between the different diagnoses is not particularly different, and many clinicians would have difficulty distinguishing between the separate diagnoses based on these lab values alone. They would normally request further tests such as imaging or biopsy to clarify the diagnosis. As demonstrated by the high probabilities above, our hybrid algorithm would be able to provide diagnostic confidence much earlier in the care pathway, possibly streamlining the path to appropriate management measures.

## DISCUSSION

Liver transplant recipients often develop elevated liver enzymes post-transplant, indicating potential graft issues^3^. Upon detection, further diagnostic steps like imaging and biopsy are pursued, though they carry risks and may lead to delays in therapy. With no reliable noninvasive tools available, a probabilistic diagnostic ranking system could expedite treatment decisions and mitigate risks, as presented in our study. Our methodology using a multi-class neural network gave the best performance in predicting each individual category of diagnosis as well as in terms of overall AUC of 0.902 as compared to the conventional machine learning approaches. We also observed no overlap in terms of the 95% confidence interval of our neural network approach [95% C.I.: 0.884, 0.919] versus the second-best performing approach of Random Forest [95% C.I: 0.812, 0.839] validating the improvement provided by our proposed methodology.

The implementation analysis (refer to Section 3) underscored the potential superiority of our ML model over the clinical judgment of clinicians. However, it also revealed an opportunity to refine our misclassification outputs through this analysis. By integrating clinician-based probabilities into our ML model, we imposed logical constraints that reflect the underlying principles of medical diagnosis. These constraints serve as regularization mechanisms, guiding the model to focus on relevant features and preventing it from overfitting to noisy or irrelevant data. As a result, the model’s predictions become more robust and reliable, increasing our overall AUC from 0.885 (ML model only) to 0.902 (ML model + clinical expertise), as they are aligned with established medical knowledge. Ultimately, the synergy between ML models and clinician expertise holds tremendous potential to optimize patient outcomes. The external validation strengthens the model performance with an AUC of 0.934, showing robustness of our approach on a completely unseen dataset.

Reviews of existing studies show that various ML algorithms, including neural networks, have been used in the context of liver disease and transplantation^12^. However, studies that focus on using ML to distinguish between various graft-related complications solely from demographic or biochemical parameters are limited^12^. For example, a study by Hughes et al from 2001 found that an artificial neural network trained on data from 117 patients with biopsies could predict the presence of ACR with an AUC of 0.902^18^. This study is limited by its small sample size and the fact that the algorithm can only diagnose one disease state – ACR. Other such examples specific to post-transplant complications include predicting recurrence of primary disease, patient and graft survival, acute kidney injury and HCC recurrence^21^. Most studies examining ML in the context of liver disease are those that automate diagnosis via image analysis of histopathologic slides^22,23^. This highlights our algorithm’s strength as the first demonstration of a neural network that can distinguish between multiple disease states based on demographic and laboratory data alone.

Neural Networks are usually perceived as black boxes wherein they improve predictive performance but are unable to provide the clinical variables driving the predictive ability. To enable interpretability of our ML modeling, we explored two avenues: Firstly, through integrated gradient methodology, we were able to identify the most important clinical variables relevant to the diagnosis of each disease state. For example, important clinical variables for ACR included elevation in ALT, AST and ALP elevation, which are well known to occur in the setting of ACR^24^. ACR was also associated with recipient age, donor age and creatinine which are all known to be associated with ACR^25^. Recurrent AIH was most associated with recipient age, consistent with a recent large study that implicated younger recipient age as a risk factor for recurrence^26^. There was also a stronger association with cyclosporine than tacrolimus use, consistent with studies of the European liver transplant registry that found cyclosporine use after liver transplant for AIH predicted worse survival when compared to tacrolimus use^27^. Expectedly, post-LT biliary complications were most associated with ALP and total bilirubin, commonly regarded as the most important biochemical parameters for diagnosis of biliary obstruction^28^. Lastly, recurrent MASH was associated with most of the clinical variables used for analysis, including hemoglobin, ALP, CNI use, and creatinine. MASH is often associated with chronic kidney disease before and after transplant, explaining the importance of creatinine in our algorithm^29^.

Secondly, through probability modeling, we were able to generate risk of graft etiology for each patient, for example, our randomly chosen patient with ACR had lab values which could be associated with various diagnoses other than ACR, including biliary obstruction or recurrent autoimmune hepatitis. Despite these lab values, our algorithm determined the probability of ACR to be 81%. This shows that our algorithm could allow prompt initiation of ACR treatment, potentially expediting management, reducing resource use, patient morbidity, and preserving graft and overall survival.

We acknowledge that the sample size for HCV-related graft injury was larger in our primary dataset, reflecting the overall cohort collected from 1992 onwards. However, recognizing that the clinical landscape has evolved and HCV is no longer a predominant cause of graft injury, we have retrained our neural network on stratified samples, differentiating between patients with HCV and non-HCV as the primary transplant indication. Our multiclass NN model demonstrates comparable performance to the original dataset, as presented in Supplementary Table 7. This finding suggests that the model’s predictive power remains robust, even as the prevalence of HCV in modern transplant populations declines.

We are also working towards creating a clinician-facing interactive dashboard for our proposed hybrid ML modeling (as shown in Figure 4(b)), where clinicians can load patient covariates such as laboratory data (blood work, liver enzymes, etc.), demographic data and clinical data (data on cholangitis, diabetes etc.) directly from the patient’s digital record, and run our ML model along with their feedback as input in the browser to return a probability of the patient being classified into one of the six post LT complications and furthermore, also get a list of the top clinical features instrumental in predicting the post LT complication in the patient. This dashboard will have the potential to inform the clinician to proactively monitor the most important clinical features in the patient making our ML approach more clinically relevant and useful.

We acknowledge some limitations of our study such as the exclusion of biopsies with dual diagnoses, potentially limiting generalizability at the current time. Additionally, we did not review the pathologies themselves and used solely the pathology report for diagnosis. Although, we might have missed some undiagnosed post-transplant complications, our sample was large and representative enough to offer sound observations. Many patients are treated empirically for mild rejection, without a liver biopsy having been performed. Regardless, we believe this work will serve as a strong basis for further development and future studies in this area. We also acknowledge the limited sample size in the external validation cohort. To address this, we applied bootstrapping and internal cross-validation to generate confidence intervals for our performance metrics and conducted sensitivity analyses, which demonstrated consistent trends across all six categories. Future validation using larger, independent datasets will be a key area for further exploration.

## CONCLUSIONS

In conclusion, our hybrid multi-class neural network model, GraftIQ, demonstrates promising potential for non-invasive diagnosis of graft pathology in liver transplant recipients. By combining clinical expertise with efficient deep learning methodologies, we offer a robust framework for accurate diagnosis, potentially reducing the reliance on invasive procedures and improving patient outcomes. If validated and implemented clinically, we believe that this method has potential to decrease time to diagnosis, dependency on liver biopsy and lead to earlier therapeutic interventions that will improve graft and patient survival over time.

## Data Availability

All data produced in the present study are available upon reasonable request to the authors

## List of Abbreviations

ACR: Acute cellular rejection
AIH: alloimmune/autoimmune hepatitis
ALP: Alkaline Phosphatase
ALT: Alanine Aminotransferase
AST: Aspartate Aminotransferase
BMI: Body Mass Index
BO: Biliary obstruction
CMV: Cytomegalovirus
EBV: Epstein Barr virus
HCC: Hepatocellular Carcinoma
HCV: Hepatitis C virus
IG: Integrated Gradient
LT: Liver transplantation
LTR: Liver transplant recipients
MELD: Model For End-Stage Liver Disease
(MASH): metabolic-associated steatohepatitis
ML: Machine learning
(NN): Neural Network
UHN: University Health Network.

## Acknowledgments

We wish to thank Mary Grace Wong and Shruti Misra for helping to review the pathology reports.

## Notes

**Conflict of Interest:** The authors have no conflicts of interest to disclose.

**Financial Support:** Supported by a Canadian Society of Transplantation grant, American Society of Transplant (AST) grant, Canadian Institutes of Health Research’s (CIHR) grant to MB. Grants not specifically for this unfunded study. The content is solely the responsibility of the author. This study was not funded by industry.

### Competing Interest Statement

The authors have declared no competing interest.

### Funding Statement

This study was funded by a Canadian Society of Transplantation grant, American Society of Transplant (AST) grant, Canadian Institutes of Health Research's (CIHR) grant to MB. Grants not specifically for this unfunded study. The content is solely the responsibility of the author. This study was not funded by industry.

### Author Declarations

Ethics committee/IRB of University Health Network gave ethical approval for this work. This study was approved by the Research Ethics Board at UHN (REB study # 21-6170). Since data was retrieved from medical records, exemption from informed consent was granted by the REB committee.

