## Supplementary figures and tables for "GraftIQ: A Hybrid Multi-Class Neural Network Integrating Clinical Insight for Multi-Outcome Prediction in Liver Transplant Recipients"

Table of Content Page

Table of contents and List of Authors ---------- 1
Supplementary Table1 ---------- 2
Supplementary Table2 ---------- 4
Supplementary Table3 ---------- 4
Supplementary Table4 ---------- 5
Supplementary Table5 ---------- 5
Supplementary Table6 ---------- 6
Supplementary Table7 ---------- 7
Supplementary Figure 1 ---------- 8
Supplementary Figure 2 ---------- 9
Supplementary Figure 3 ----------10
Supplementary Figure 4 ----------11
Supplementary Figure 5 ----------12

**Supplementary Table1:** Primary Indication of Transplant in Subjects with at least one biopsy information

| Covariate | N = 1791 |
|---|---|
| **Transplant Primary Indication** | |
| Acute & Subacute Necrosis of Liver | 1 (0) |
| Acute Fulminant Hepatic Failure – Viral – Other | 1 (0) |
| Acute Intermittent Porphyria | 1 (0) |
| Acute on Chronic Liver Failure | 1 (0) |
| Alagille's Syndrome | 3 (0) |
| Alcoholic Cirrhosis Of Liver | 201 (11) |
| ALD Protocol / Alcoholic Cirrhosis Of Liver | 10 (1) |
| Alpha I Anti-trypsin Deficiency | 6 (0) |
| Alpha I Antitrypsin Deficiency | 14 (1) |
| Amyloidosis | 6 (0) |
| Autoimmune Active Hepatitis | 46 (3) |
| Autoimmune Hepatitis | 12 (1) |
| Biliary Atresia | 2 (0) |
| Budd-Chiari Syndrome | 9 (1) |
| Caroli'S Disease | 2 (0) |
| Cirrhosis | 1 (0) |
| Cirrhosis-Other | 1 (0) |
| Cirrhosis, Unknown Etiology | 44 (3) |
| Cryptogenic Cirrhosis | 6 (0) |
| Cystic Fibrosis | 4 (0) |
| Cystic Fibrosis-Cepacia Negative | 3 (0) |
| Drug Induced Hepatitis | 1 (0) |
| Epithelioid Hemangioendothelioma | 1 (0) |
| Familial Cholestasis | 1 (0) |
| Fulminant Hepatic Failure | 44 (3) |
| Fulminant hepatic failure - acetaminophen | 3 (0) |
| Fulminant hepatic failure - drug, other | 1 (0) |
| Fulminant hepatitis B | 1 (0) |
| Giant Cell Hepatitis | 1 (0) |
| Glycogen Storage Disease | 2 (0) |
| Haemochromatosis | 4 (0) |
| Hemangioma, Any Site | 1 (0) |
| Hepatitis - Unknown | 3 (0) |
| Hepatitis B | 134 (8) |
| Hepatitis C | 601 (34) |
| Hepatitis C - Hepatitis C virus (HCV) RNA negative | 53 (3) |
| Hepatitis C - Post Tx | 1 (0) |
| Hepatitis C - RNA Pos | 57 (3) |
| Hepatitis G | 1 (0) |

| | |
|---|---|
| Hepatitis Non A/NonB | 2 (0) |
| Hepatitis, Unspecified | 1 (0) |
| Hepatopulmonary Syndrome | 4 (0) |
| Hepatorenal syndrome | 1 (0) |
| Idiopathic Fibrosing Alveolitis | 1 (0) |
| Malignancy - cholangiocarcinoma | 3 (0) |
| Malignancy - Fibrolamellar hepatoma | 1 (0) |
| Malignancy - Hepatoma | 15 (1) |
| Maple Syrup Urine Disease | 1 (0) |
| Mesenteric Thrombosis | 1 (0) |
| Metabolic Disorder – Hemochromatosis | 4 (0) |
| Metabolic Disorder – Wilson’S Disease | 1 (0) |
| Mitochondrial Disease | 1 (0) |
| Nonalcoholic steatohepatitis (NASH) | 121 (7) |
| Other Metabolics | 3 (0) |
| Polycystic | 1 (0) |
| Polycystic Liver Disease | 4 (0) |
| Primary Biliary Cholangitis | 90 (5) |
| Primary Sclerosing Cholangitis | 141 (8) |
| Re-Tx: Chronic Biliary Obstruction | 11 (1) |
| Re-Tx: Chronic Rejection | 1 (0) |
| Re-Tx: Recurrent Disease | 6 (0) |
| Re-Tx: Vessel Thrombosis | 2 (0) |
| Recurrent Cholangitis | 1 (0) |
| Retransplant | 2 (0) |
| Retransplantation | 31 (2) |
| Sarcoidosis | 1 (0) |
| Sickle Cell Crisis | 1 (0) |
| Sickle cell disease | 2 (0) |
| Sub-Acute Fulminant | 1 (0) |
| Subacute Fulminant Hepatic Failure – Viral – Other | 1 (0) |
| Tylenol (acetaminophen) overdose | 1 (0) |
| Wilson’S Disease | 8 (0) |
| Missing | **42** |

**Supplementary Table 2:** Tuning alpha and beta values to obtain the optimal alpha and beta values for bayesian fusion of clinical expertise and ML tool predictions.

| alpha | beta | Overall AUC |
|---|---|---|
| 0.1 | 0.9 | 0.887 |
| 0.2 | 0.8 | 0.902 |
| 0.3 | 0.7 | 0.897 |
| 0.4 | 0.6 | 0.866 |

**Supplementary Table 3:** Evaluation results using mean Area Under the Curve (AUC) metric and 95% Confidence Intervals (CI) on each diagnosis category in the external validation dataset using the proposed GraftIQ model

| Categories | AUC [95% CI] |
|---|---|
| AIH | 0.962 [0.949, 0.974] |
| BO | 0.945 [0.933, 0.956] |
| Congestion | 0.923 [0.909, 0.935] |
| HCV | 0.922 [0.909, 0.933] |
| ACR | 0.898 [0.886, 0.908] |
| MASH | 0.951 [0.939, 0.964] |

**Supplementary Table 4:** Hyperparameter optimization for multiclass Neural Network algorithm

| Hyperparameter | Value | Search Range |
|---|---|---|
| Learning Rate | 0.005 | 0.1, 0.01, 0.005, 0.001, 0.0005 |
| Optimization solver | Adam | Stochastic Gradient Descent, Adam |
| Feature Scaling | Standard | Min-Max and Standard scaler |
| Number of Layers | 3 | 1,3,5,7 |
| Hidden units/ layer | 24 | 12,24,48,96,192 |
| Number of epochs | 400 | 100-600 |
| Dropout rate | 0.5 | 0.3,0.4,0.5,0.6,0.7,0.8 |

**Supplementary Table 5:** Hyperparameter optimization for conventional Machine Learning Algorithms used for comparison to our proposed methodology.

| Approaches | Parameter | Value | Search Range |
|---|---|---|---|
| Random Forest | No. of Trees | 400 | [10, 500] |
| | Mtry (No. of columns to randomly select at each level) | 380 | [50,400] |
| SVM Classifier | C | 1 | [0.1, 100] |
| | Kernel | sigmoid | 'linear', 'poly', 'rbf', 'sigmoid' |
| Lasso Regression | alpha | 0.005 | [0.001, 0.1] |
| Ridge Regression | alpha | 0.005 | [0.001, 0.1] |
| Logistic Regression | C | 0.001 | [0.1,100] |

**Supplementary Table 6:** Input variables and their missing rates

| Variables | Definition of variables | Missing values (N=1791) | % Missing |
|---|---|---|---|
| Recipient gender | Sex (Recipient) | 0 | 0 |
| Recipient age | Age (Recipient) | 0 | 0 |
| WBC | White Blood cell (WBC) | 73 | 3.8 |
| platelets | Platelets | 79 | 4.1 |
| Creatinine | Serum Creatinine | 75 | 3.9 |
| INR | International normalized ratio | 100 | 5.3 |
| Albumin | Serum Albumin | 407 | 21.5 |
| Sodium | Serum Sodium (Na) | 71 | 3.7 |
| Tacrolimus serum level | Tacrolimus serum level | 397 | 21 |
| Cyclosporine serum level | Cyclosporine serum level | 397 | 21 |
| Hemoglobin | Hemoglobin | 72 | 3.8 |
| Bilirubin | Serum Total bilirubin | 82 | 4.3 |
| AST | Aspartate aminotransferase (AST) | 72 | 3.8 |
| ALT | Alanine aminotransferase (ALT) | 60 | 3.1 |
| ALP | Alkaline phosphatase (ALP) | 67 | 3.5 |
| MELD score | MELD score | 0 | 0 |
| Transplant primary indication | Primary indication for transplant | 0 | 0 |
| Donor Type (LDLT vs DDLT) | Living vs deceased donor | 0 | 0 |
| Weight | Weight (kg) | 0 | 0 |
| BMI | Body Mass Index (BMI) | 0 | 0 |
| Diabetes Mellitus | Diabetes Mellitus at transplant (Yes/ No) | 0 | 0 |
| Hypertension | Hypertension at transplant (Yes/No) | 0 | 0 |
| Dyslipidemia | Dyslipidemia at transplant (Yes/No) | 0 | 0 |

**Subgroup analysis based on primary indication for LT (HCV vs non-HCV):**

In this study, we conducted a subgroup analysis based on the primary indication for liver transplantation (LT), specifically comparing patients transplanted for hepatitis C virus (HCV)-related liver disease to those with non-HCV etiologies. The purpose of this analysis was to assess the impact of the primary indication for transplant on the prediction of graft injury categories.

For the non-HCV cohort, we included 1,322 patients with acute cellular rejection (ACR), 102 with alloimmune hepatitis (AIH), 333 with biliary obstruction (BO), 112 with congestion, and 129 with metabolic associated steatohepatitis (MASH). As there was no HCV graft injury category for patients in the non-HCV primary transplant indication group, we retrained our model to test its performance on five graft injury categories (ACR, AIH, BO, congestion, and MASH) for this subgroup. In contrast, the HCV cohort comprised 566 patients with ACR, 82 with AIH, 147 with BO, 51 with congestion, 2,181 with recurrent HCV, and 119 with MASH. This stratification allowed us to compare the model's performance across different graft injury categories for both HCV and non-HCV patients, ensuring that our model remains robust and generalizable to modern transplant populations, where HCV is less prevalent.

**Supplementary Table 7: Comparison of AUC values for the multiclass neural network (NN)-based machine learning (ML) model in predicting graft injury categories in HCV and non-HCV liver transplant recipients.** The table shows the model's performance across six categories: Acute Cellular Rejection (ACR), Alloimmune Hepatitis (AIH), Biliary Obstruction (BO), Congestion, HCV recurrence, and Metabolic Associated Steatohepatitis (MASH). AUC values demonstrate consistent model performance across both HCV and non-HCV cohorts, indicating the model's robustness.

| Category | AUC multiclass NN based ML model | AUC multiclass NN based ML model |
|---|---|---|
| | Primary indication of transplant HCV | Primary indication of transplant Non-HCV |
| ACR | 0.781 | 0.769 |
| AIH | 0.919 | 0.907 |
| BO | 0.908 | 0.896 |
| Congestion | 0.915 | 0.903 |
| HCV | 0.864 | - |
| MASH | 0.910 | 0.902 |

**Supplementary Figure 1:** Important variables for predicting diagnosis in AIH category

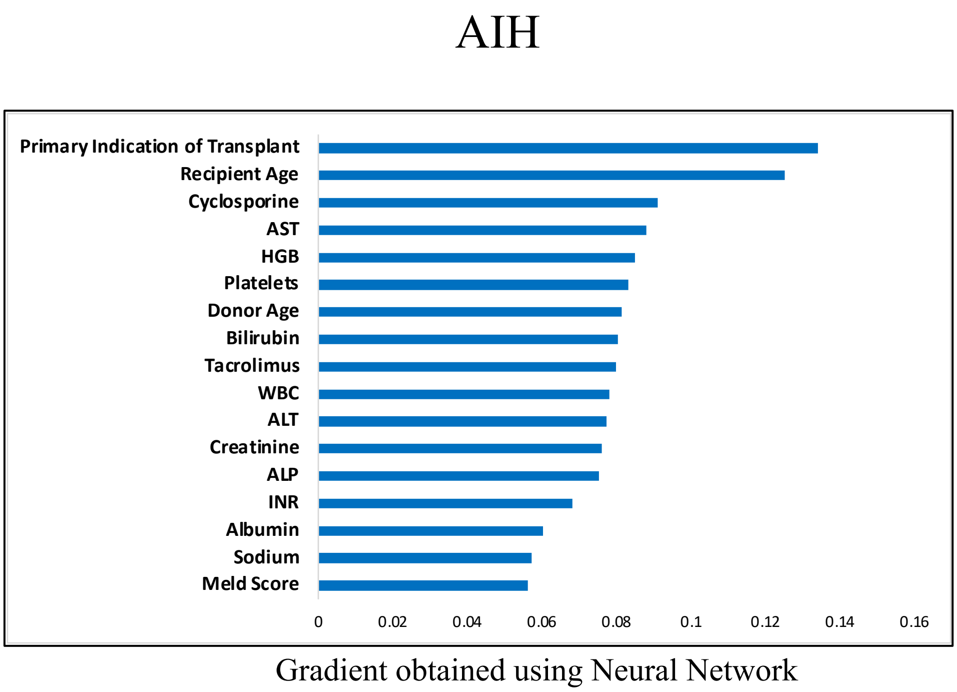

**Supplementary Figure 2:** Important variables for predicting diagnosis in congestion category

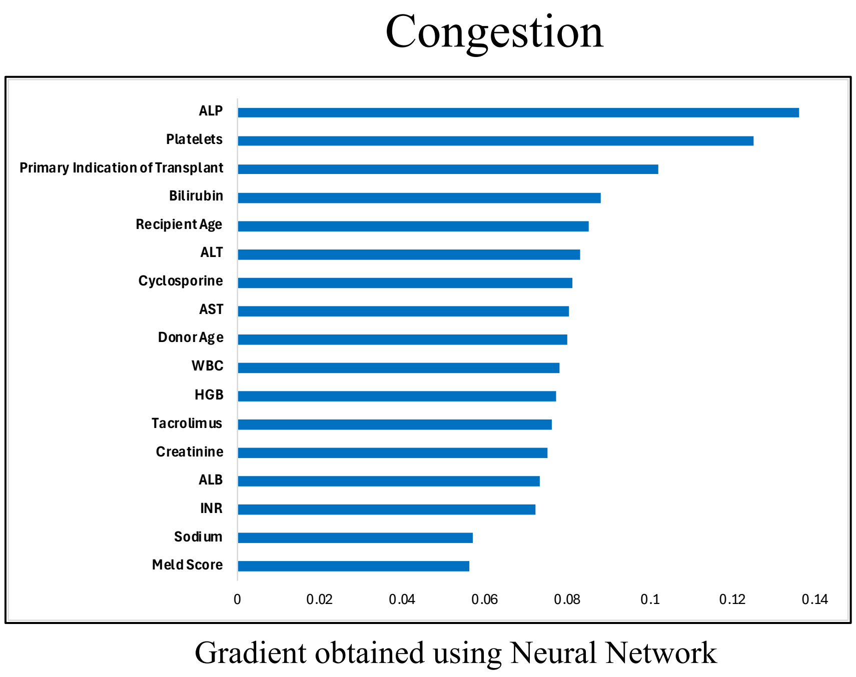

**Supplementary Figure 3:** Important variables for predicting diagnosis in BO category

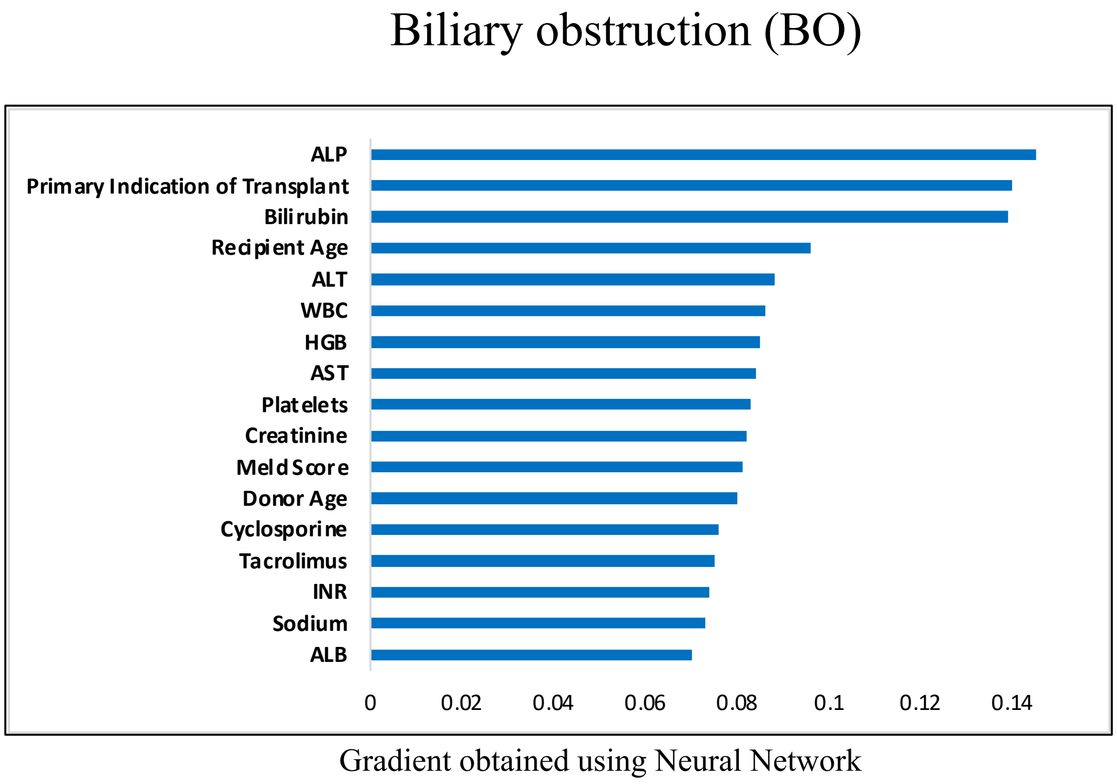

**Supplementary Figure 4:** Important variables for predicting diagnosis in HCV category

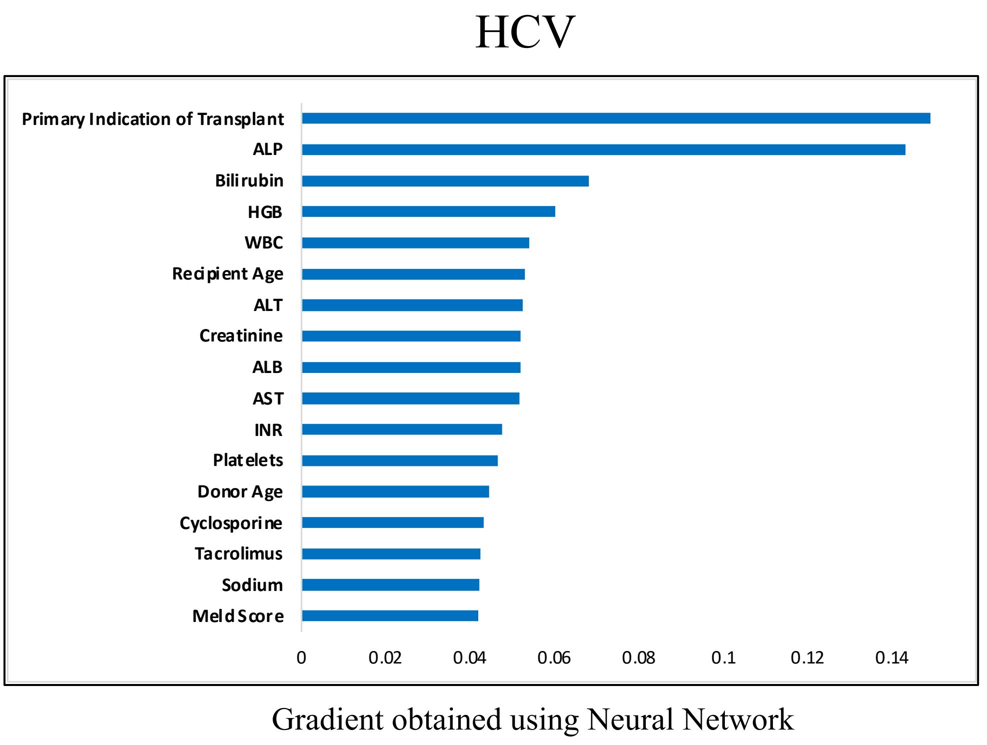

**Supplementary Figure 5:** Important variables for predicting diagnosis in MASH category

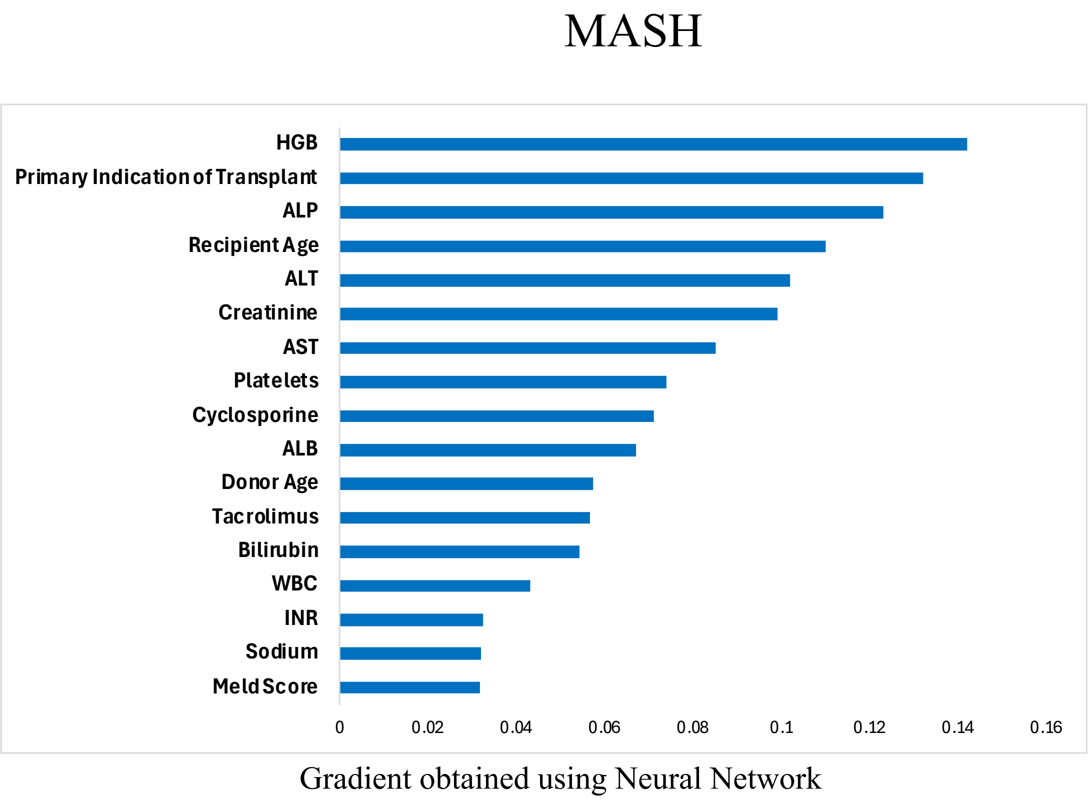
